# NRX-101 (D-cycloserine plus lurasidone) vs. lurasidone for the maintenance of initial stabilization after ketamine in patients with severe bipolar depression with acute suicidal ideation and behavior: A randomized prospective phase 2 trial

**DOI:** 10.1101/2022.08.11.22278658

**Authors:** Andrew Nierenberg, Philip Lavin, Daniel C. Javitt, Richard Shelton, Sanjay Matthew, Robert A Besthof, Jonathan C. Javitt

## Abstract

**Objectives:** We tested the hypothesis that after initial improvement with intravenous ketamine in patients with bipolar disorder (BD) with severe depression and acute suicidal thinking or behavior, a fixed-dose combination of oral D-cycloserine (DCS) and lurasidone (NRX-101) can maintain improvement more effectively lurasidone alone.

**Methods:** This was a multi-center, double-blind, two-stage, parallel randomized trial. Adult BD patients with depression and suicidal ideation or behavior were infused with ketamine or saline (Stage 1); those who improved were randomized to a fixed dose combination of DCS and lurasidone vs. lurasidone alone (Stage 2) to maintain the improvement achieved in Stage 1. Depression was measured by the Montgomery Åsberg Depression Rating Scale (MADRS) and suicidal thinking and behavior was measured by the Columbia Suicide Severity Rating Scale (C-SSRS); global improvement was measured by the clinical global severity scale (CGI-S).

**Results:** Thirty-seven patients were screened and 22 were enrolled, randomized, and treated. All 22 patients treated in Stage 1 (17 with ketamine and 5 with saline) were enrolled into Stage 2 and 11 completed the study. The fixed dose combination of DCS and lurasidone was significantly more effective than lurasidone alone in maintaining improvement in depression (MADRS LMS Δ-7.7; p=.03) and reducing suicidal ideation, as measured by C-SSRS (Δ-1.5; p=0.02) and by CGI-SS (Δ-2.9; p=0.03), and with a non-statistically significant decrease in depressive relapse (0% vs. 40%; p=0.07). This sequential treatment regimen did not cause any significant safety events and demonstrated improvements in patient reported side effects.

**Conclusions:** Sequential treatment of a single infusion of ketamine followed by NRX-101 maintenance is a promising therapeutic approach for reducing depression and suicidal ideation in patients with bipolar depression who require hospitalization due to acute suicidal ideation and behavior. On the basis of these findings, Breakthrough Therapy Designation was awarded and a Special Protocol Agreement was granted by the FDA for a registrational trial.

## INTRODUCTION

Bipolar disorder (BD) has a lifetime prevalence of 4.4% in adults in the United States and the risk of acute suicidal ideation and behavior (ASIB) is uniquely high in patients during bipolar depressive episodes.^1,2^ It is estimated that up to 50% of individuals with BD may have suicidal behavior over their lifetime^3^, and between 11% and 19.9%^4^ may die from suicide. Patients with BD are 20-30 times more likely to have suicidal behavior than the general population^5^ and the overall rate of death by suicide among patients with BD is approximately 10-fold greater than that of the general population.^6^

Despite its lethal characteristics, there is no approved pharmacologic treatment for patients with bipolar depression in the presence of ASIB. Electroconvulsive therapy (ECT), often combined with inpatient psychiatric care, remains the only FDA-approved treatment for patients with ASIB in bipolar depression. Several combined dopamine D2 receptor and serotonin 5-HT_2A_ receptor antagonists, such as olanzapine/fluoxetine, quetiapine, lurasidone, cariprazine, and lumateparone, have demonstrated efficacy in treating bipolar depression, however, none has been shown to decrease ASIB and all have an FDA-mandated warning regarding a potential to increase the risk of suicide.^7,8^ Indeed, patients with suicidal ideation are routinely excluded from clinical trials of experimental anti-depressive agents (e.g. clinicaltrials.gov NCT-04019704). ^9^ Accordingly, ASIB in bipolar depression must frequently be treated with voluntary or involuntary hospitalization under highly supervised conditions with untested medications and, less frequently with ECT, and represents a major unmet medical need. Furthermore, the risk of suicide after a hospitalization for psychiatric conditions can be elevated.^10^

Functional antagonists of the N-methyl D-aspartate Receptor (NMDAR) were first hypothesized to have antidepressant effects by Trullas and Skolnick. ^11^ The discovery that ketamine has a rapid and profound effect on depression and suicidality ^12^ has led to broad recognition that glutamate system and the antagonists may also play an important role in depression and suicidality. Ketamine has since been shown in multiple randomized clinical trials to induce nearly immediate remission from depressive symptoms as well as from suicidal ideation. The improvements in ASIB appear to be distinct from improvements in depression.^13,14^ However, the clinical effect has been demonstrated to diminish very quickly, e.g., three days post-intravenous dosing^15,16^ and 2 days post-intranasal dosing.^17^ Moreover, ketamine is an addictive, controlled substance that frequently causes hallucination, has known neurotoxicity, and exhibits abuse potential. The American Psychiatric Association and other expert bodies have expressed concern about the repeated use of ketamine for the treatment of depression and/or suicidality.

D-cycloserine (DCS) is a broad-spectrum antibiotic approved for the treatment of tuberculosis that has been used in millions of individuals without report of significant safety concerns or abuse potential. DCS was serendipitously found to have antidepressant effects by Crane, ^18^ which were subsequently confirmed in a placebo-controlled trial. ^19^ However, psychotomimetic side-effects noted by Crane led investigators to focus on iproniazide and imipramine as first-generation oral antidepressants.

Given the known role of NMDAR in both psychogenic^20^ and antidepressant effects of NMDAR^21^, the antidepressant effect of DCS was subsequently attributed to it partial agonist effect at the glycine site of the NMDA receptor.^22^ DCS was shown to have potential antidepressant effects on rodent forced swim test by Lopes and coworkers.^23^ In humans, DCS primarily manifests NMDA agonist effects at doses below 50 mg/d whereas it primarily manifests NMDA antagonist effects at doses of >500 mg/d. For this reason, numerous trials with low-dose DCS in depression and PTSD have failed to demonstrate clinical effect. DCS at a target dose of >500mg/d has been shown to have a clinical antidepressant effect when administered to patients with treatment resistant depression added to SSRI antidepressants, without hallucinogenic side effects.^24^ DCS sustained the antidepressant effect of ketamine in an open label trial in patients with bipolar depression who were resistant to currently approved pharmacological treatments. ^25^ DCS has further been demonstrated to maintain remission from suicidality after ketamine infusion, an effect that is more pronounced among patients with bipolar depression.^26^

DCS is a potentially attractive oral antidepressant candidate because unlike direct NMDA channel blocking agents, it has shown no potential for neurotoxicity or addiction. Moreover, in non-clinical models it has been shown to decrease akathisia induced by 5-HT_2A_ targeted drugs, including atypical antipsychotic agents such as lurasidone.^27^

In this study, we tested the hypothesis that a fixed-dose combination of oral DCS and lurasidone (NRX-101) could better maintain initial stabilization from severe depression and ASIB in acutely depressed BD patients than could lurasidone alone (i.e. an active comparator) following improvement with a single infusion of ketamine.

## PATIENTS and METHODS

### Study design

The study was conducted in accordance with the ethical principles as set out in the Declaration of Helsinki and its amendments and in compliance with the approved protocol of the United States Food and Drug Administration (FDA). The protocol and its associated Informed Consent Agreement were reviewed and approved by the central Institutional Review Board (IRB) of this study Schulman IRB (now Advarra, Inc). Participants were evaluated to ensure that they were capable of understanding the nature of this study and its potential risks, discomforts, and benefits. All participants provided written informed consent. The study was conducted at one academic site (UAB, Birmingham) and two community hospital locations (see clinicaltrials.gov).

This was a multi-center, double-blind, two-stage, parallel randomized trial to evaluate the efficacy of ketamine, compared to saline, in the rapid improvement of patients with severe BD and ASIB (Stage 1) and the efficacy of NRX-101, compared to lurasidone, in maintaining the improvement achieved in Stage 1 (Stage 2). There were separate randomizations for Stage 1 and Stage 2.

This study was originally designed for a larger sample size but was terminated prior to full enrollment to accommodate a Special Protocol Agreement (SPA) with FDA wherein the primary endpoint scale was changed from the Bipolar Inventory of Symptoms Scale (BISS)-derived Montgomery-Åsberg Depression Rating Scale (MADRS) (BDM) to the traditional MADRS 10 scale. This request was made by FDA to facilitate post-hoc comparison of NRX-101 results with placebo results already on file at FDA. The FDA additionally requested a separation of Stage 1 and Stage 2 for the SPA protocol. This study enrolled, randomized, and treated 22 patients prior to its early termination.

### Recruitment and eligibility

The study was conducted at one academic and two non-academic research sites in the US. Key eligibility criteria included adults 18 to 65 years of age diagnosed with BD (according to the criteria defined in the Fifth Edition of the Diagnostic and Statistical Manual of Mental Disorder with a score of ≥ 20 on the BDM) and suicidal ideation or behavior [an answer of “Yes” to item 4 or item 5 on the Columbia Suicide Severity Rating Scale (C-SSRS)] who were able to understand and provide informed consent, deemed likely to comply with study protocol and communicate adverse events (AEs) and other clinically important information and agreed to be hospitalized to complete screening and initiate experimental treatment. Eligible participants were required to have resided in a stable living situation, had an identified reliable informant, and were in good general health. In addition, females were required to have a status of non–childbearing potential or use of an acceptable form of birth control. Concurrent psychotherapy and hypnotic therapy were allowed if the therapy had been stable for at least three months (for psychotherapy) or four weeks (for hypnotic therapy) prior to screening and if the therapy was expected to remain stable during the study.

Key exclusion criteria included females who were pregnant, breastfeeding, or of childbearing potential and were unwilling to use one of the specified forms of birth control during the study; individuals who had a moderate or severe substance use disorder within the 12 months prior to screening, had a lifetime history of phencyclidine or ketamine drug use, had failed use of ketamine for depression, had any history of psychotic symptoms when not in an acute bipolar mood episode, had any major psychiatric disorder that was clinically predominant to BD at screening or with BD as the primary or secondary focus of treatment at any time within six months prior to screening, or had dementia, delirium, amnestic, or any other cognitive disorder; individuals who had a history or a current episode of hypertension or a current episode of cardiac abnormality. Additional exclusion criteria are shown on clinicaltrials.gov 1.

### Study intervention

The Investigator, Patient, and Study Staff were blinded to treatment assignment, except for the unblinded pharmacist who prepared the Stage 1. Packaging and labelling of the study drugs were performed to ensure blinding throughout the study by providing uniquely assigned kit numbers per randomized patient and identical appearance of NRX-101 and lurasidone comparator capsules.

For Stage 1, the experimental drug was ketamine (0.5 mg/kg ketamine hydrochloride injection purchased from PAR Sterile Products, 870 Parkdale Rd., Rochester, MI 48307), which was administered once through intravenous infusion over a 40-minute duration, and the reference was normal saline (0.9% sodium chloride 100 mL). For Stage 2, the experimental drug was a fixed dose combination of DCS and lurasidone (NRX-101: NRx Pharmaceuticals, Wilmington, DE) and the comparator was lurasidone at the same dosage as in the matched experimental drug. NRX-101 and lurasidone comparator were manufactured to Good Manufacturing Practices (GMP) standards at an FDA-inspected facility (WuXi Apptec: Shanghai, CN). NRX-101 was administered orally twice a day, starting on Day 1 or 2 depending on the timing of the response in Stage 1, and continued for 6 weeks. The total doses of DCS and lurasidone were 350 mg and 33 mg, respectively, for the first administration and were up-titrated to 950 mg DCS and 66 mg lurasidone by the 5^th^ administration. After clinical evaluation, the blinded Investigator could up-titrate the lurasidone dose two steps (to 99 and 132 mg/day) or down-titrate DCS and lurasidone two steps (DCS: 825 and 700 mg/day; lurasidone 49.5 and 33 mg/day) (Appendix Tables 2, 3, 4). Due to reports from the tuberculosis literature suggesting that high-dose DCS may alter pyridoxine, all patients were given Vitamin B6 tablets (pyridoxine, 100mg) and instructed to take one tablet daily to prevent potential side effects with the investigational drug under study.

### Randomization

Stage 1 randomization at study entry assigned patients to ketamine versus saline in a 3:1 ratio, Stage 2 randomization assigned patients to DCS plus lurasidone versus lurasidone in a 2:1 ratio. Stage 1 randomization was stratified by baseline C-SSRS score (4 vs. 5), presence vs absence of sub-syndromal hypomanic symptoms (determined by BDM score), and evidence of a suicidal event in the prior 12 months [including an attempt, an Emergency Department visit or hospitalization for suicidality or mental health encounter prompted by suicidality]. All patients with at least a minimal improvement (defined as ≥25% improvement in BDM, and C-SSRS ≤3) assessed 24 and 48 hours after infusion were offered the option to continue into Stage 2, a 42-day treatment phase.

Stage 2 randomization was stratified by the Stage 1 treatment, by the Stage 1 response [partial (25-49% CFB BDM) or full response (≥50%]), and if directly assigned to ketamine (in the event that saline infusion was declared futile by the Data Safety Monitoring Board). IWRS failures requiring manual treatment assignments were recorded.

### Treatment Compliance Monitoring

Treatment compliance was monitored using a Health Insurance Portability and Accountability Act-compliant mobile phone–based visual imaging technology, AiCure. The platform allowed for confirmation of oral dosing of investigational product and date/time of the dosing and had built-in reminders and a communication system allowed for real-time intervention in case of drug interruptions. The platform was preloaded on a smartphone provided to patients during the first visit. At no time was the phone number visible to healthcare providers or monitoring personnel. Individuals outside the clinical sites were not provided with patient names, nor were given access to patient medical records. At the end of the study, the patients returned the phones to the study site. Patients who missed one visit were allowed to make up that visit at the end of the dosing week. Patients who missed more than one visit were discontinued from the study.

### Outcome measures

The primary efficacy endpoint for the Stage 1 was the proportion of patients with a successful response, as measured by acutely diminished suicidality and improved mood (achieving C-SSRS ≤3 and an acute ≥25% improvement in depression ratings) within 48 hours. The primary efficacy endpoint for Stage 2 was the improvement in the BDM from Stage 2 baseline through Day 42 of the treatment in those patients who responded in Stage 1 using a Mixed Model Repeated Measures (MMRM) analysis without and with Last Observation Carried Forward (LOCF).

The key secondary efficacy endpoint of Stage 1 was the difference in mean change on the BDM between ketamine and saline groups assessed at Day 1 and Day 2. The planned key secondary efficacy endpoint in Stage 2 was the difference in time to relapse between the two treatment arms. Because no instances of relapse occurred in the NRX-101 group, a time-to-event analysis could not be performed and relapse rates of the two treatment groups were compared. Relapse was defined as any one of the following events during Stage 2: C-SSRS score of ≥4, BDM score ≥20, implementation of a new treatment plan, or the addition of new antidepressant drugs. Relapse was adjudicated by a three-physician Relapse Adjudication Committee on an ongoing basis because those patients determined to have relapsed were withdrawn from the study and returned to closely-monitored psychiatric care.

Akathisia is a known side effect of medications that inhibit the 5-HT_2A_ receptor and is potentially associated with increased suicidal ideation and behavior. ^28^ Nonclinical studies (D. Javitt, personal communication) have shown a significant reduction in akathisia when NMDAR-antagonist drugs are added to 5-HT_2A_ antagonists in the rodent Elevated Plus Maze assay. Accordingly, the Barnes Akathisia Rating Scale (BARS) was included as a safety measure.^29^

### Pharmacokinetics Measurements

Blood samples for pharmacokinetic (PK) analysis of ketamine were collected one hour prior to and two hours after the initial administration. Blood samples for monitoring plasma DCS and lurasidone levels were collected from all patients to verify exposure and consistency with randomization assignment. Samples for the trough determination were drawn within one hour before the drug administration on Days 14 and 42 whereas samples for the steady state Cmax determination were drawn 2 hours (±15 min) after the drug administration on Day 42. Additional PK samples were obtained from patients with serious adverse events (SAEs) in order to characterize potential relationships between AEs and plasma levels. An unblinded medical monitor reviewed these data on an ongoing basis.

### Statistical Methods

All analyses were performed using SAS statistical software, version 9.4 or later. This study was originally powered at Stage 1 (n=140) to detect a 30% difference in post-infusion response rates and powered at Stage 2 (n=72) to detect a 40% reduction in relapse rates. Because the study was stopped early to accommodate the Special Protocol Agreement with FDA, 20 patients were planned to be enrolled to confirm protocol feasibility and drug exposure. However, 22 patients were enrolled because two additional patients had consented when study termination was announced. The study, as enrolled had 80% power to detect a 7 point mean difference in primary endpoint. This magnitude of difference was observed in prior trials.^19^

There was a total of five analysis cohorts, which include Informed Consent, Stage 1 ITT22, Stage 2 ITT22, as well as Stage 1 and Stage 2 Safety Sets. Study Day 0 (pre-infusion) was the day of randomization to Stage 1, Baseline1 was the last observation prior to Stage 1 drug infusion, and Baseline2 was the last observation prior to Stage 2 drug dosing.

For the primary endpoints, descriptive statistics and Mixed Model Repeated Measures (MMRM)-based average scores and p-values were reported by visit and overall. The primary analysis was based on Last Observation Carried Forward (LOCF), as is typical in antidepressant trials.^8^ The proportion of responders and relapses between the two treatments was compared using a two-sided Fisher Exact Test. Three rating scales, Brief Psychiatric Scale (BPRS+), the Clinical Global Impression of Severity for Suicide (CGI-SS), and C-SSRS, were analyzed using paired t-tests on within treatment change and unpaired t-tests to compare change from baseline differences between treatments.

### Rater Concordance

Assuring consistency between raters within and across study sites is key to reliable measurement of primary outcome. Accordingly, all rating sessions were captured digitally and transmitted to central master raters. In order to be certified for participation in the clinical trial, raters were required to demonstrate 5 years of experience with the outcome scales used in the clinical trial and were required to demonstrate 80% concordance with master raters on the standardized training set.

## RESULTS

### Baseline Characteristics and Disposition of patients

Thirty-seven patients were screened in this study, 22 of whom were eligible for enrollment and all were randomized and received treatment (Figure 1). All 22 patients treated in Stage 1 (17 with ketamine and 5 with saline) were enrolled into Stage 2, 11 (50%) completed the study whereas the other 11 (50%) discontinued from the study. No patients were lost to follow-up.

**Figure 1:**
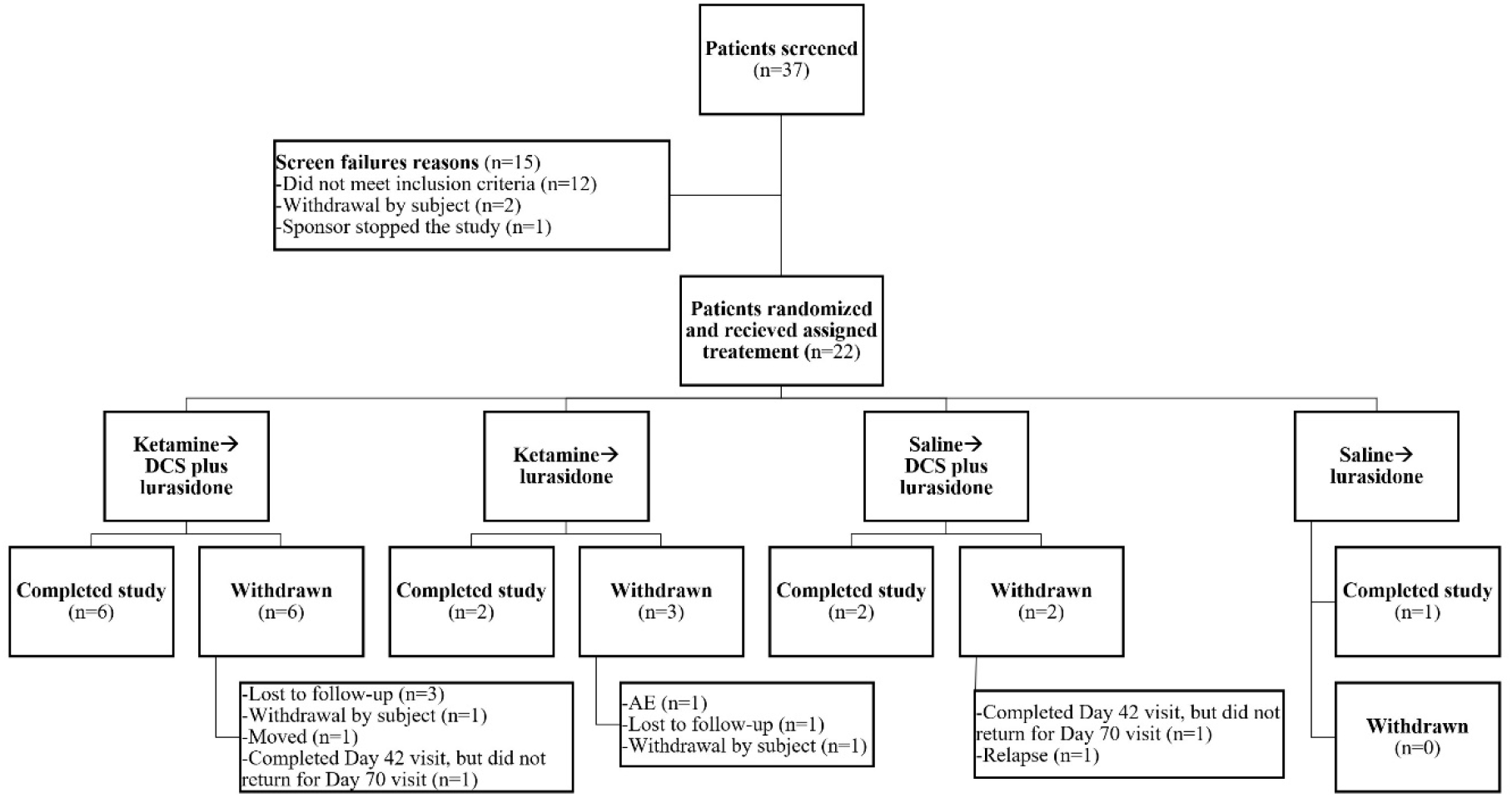
CONSORT Diagram of Screened and Enrolled Participants

Both Stage 1 and Stage 2 Safety populations consisted of all 22 patients who received a study treatment. The majority of patients in the Safety Population were Caucasian [ketamine, n=16 (94.1%); saline, n=3 (60%)], the remaining patients were Black or African American, and were male [ketamine, n=13 (76.5%); saline, n=3 (60%)]. The mean age of patients was 39.5 years (Table 1).

**Table 1.** Baseline Characteristics, and Stratification Factors.

| Parameter | Result | Saline<br>(N=5) | Ketamine<br>(N=17) | ketamine |  | saline |  | All Patients<br>(N=22) |
| --- | --- | --- | --- | --- | --- | --- | --- | --- |
|  |  |  |  | DCS plus<br>lurasidone<br>(N=12) | lurasidone<br>(N=5) | DCS plus<br>lurasidone<br>(N=4) | lurasidone<br>(N=1) |  |
| Age | Mean | 43.6 | 38.4 | 33.9 | 49.0 | 41.0 | 54.0 | 39.5 |
|  | Median | 46.0 | 42.0 | 30.0 | 45.0 | 42.0 | 54.0 | 42.5 |
|  | Std Dev | 9.63 | 11.90 | 10.47 | 8.03 | 8.87 | 0 | 11.43 |
|  | Min, Max | 30, 54 | 19, 60 | 19, 48 | 42, 60 | 30, 50 | 54, 54 | 19, 60 |
|  | 95% CI | (31.6, 55.6) | (32.2, 44.5) | (27.3, 40.6) | (39.0, 59.0) | (26.9, 55.1) | 0 | (34.5, 44.6) |
| Sex [n (%)] | Male | 3 (60.0) | 13 (76.5) | 9 (75.0) | 4 (80.0) | 2 (50.0) | 1 (100) | 16 (72.7) |
|  | Female | 2 (40.0) | 4 (23.5) | 3 (25.0) | 1 (20.0) | 2 (50.0) | 0 | 6 (27.3) |
| Race [n (%)] | White | 3 (60.0) | 16 (94.1) | 11 (91.7) | 5 (100) | 3 (75.0) | 0 | 19 (86.4) |
|  | Black or<br>African-American | 2 (40.0) | 1 (5.9) | 1 (8.3) | 0 | 1 (25.0) | 1 (100) | 3 (13.6) |
| Ethnicity [n (%)] | Hispanic | 0 | 0 | 0 | 0 | 0 | 0 | 0 |
|  | Non-Hispanic | 5 (100) | 17 (100) | 12 (100) | 5 (100) | 4 (100) | 1 (100) | 22 (100) |
| Baseline C-SSRS<br>Score [n (%)]* | 1 | 0 | 1 (5.9) | 1 (8.3) | 0 | 0 | 0 | 1 (4.5) |
|  | 4 | 0 | 2 (11.8) | 2 (16.7) | 0 | 0 | 0 | 2 (9.1) |
|  | 5 | 5 (100) | 14 8 (2.4) | 9 (75.0) | 5 (100) | 4 (100) | 1 (100) | 19 (86.4) |
| Hypomanic<br>symptoms (BISS) | None | 4 (80.0) | 13 (76.5) | 9 (75.0) | 4 (80.0) | 3 (75.0) | 1 (100) | 17 (77.3) |
|  | Any | 1 (20.0) | 4 (23.5) | 3 (25.0) | 1 (20.0) | 1 (25.0) | 0 | 5 (22.7) |
| Suicidal Event in<br>Prior 12 Months | None | 1 (20.0) | 3 (17.6) | 3 (25.0) | 0 | 1 (25.0) | 0 | 4 (18.2) |
|  | Any | 4 (80.0) | 14 (82.4) | 9 (75.0) | 5 (100) | 3 (75.0) | 1 (100) | 18 (81.8) |

BISS = Bipolar Inventory of Symptoms Scale; C-SSRS = Columbia Suicide Severity Rating Scale
\* one participant achieved spontaneous remission from suicidality prior to ketamine infusion and did not progress to stage 2 randomization.

Twenty-one of the 22 patients presented with a baseline score ≥4 on the C-SSRS [ketamine, n=16 (94.1%); Saline, n=5 (100%)]. One subject presented at study intake with C-SSRS=5 but was noted to have C-SSRS=1 at the pre-infusion assessment 24 hours later and did not progress to stage 2. The majority of patients [n=17 (77.3%)] did not present with hypomanic symptoms on the BISS [ketamine, n=13 (76.5%); Saline, n=4 (80.0%)]. Eighteen of the 22 patients had experienced at least one suicidal event in the past 12 months [ketamine, n=14 (82.4%); Saline, n=4 (80.0%)]. (Table 1)

The analysis population consisted of all 22 patients who were randomized to receive a study treatment (i.e. intent to treat). For the Stage 1 Safety and ITT22, 17 patients received ketamine and five patients received saline. Of the 17 patients who received ketamine in Stage 1, 12 patients were subsequently randomized to oral NRX-101 (ketamine → NRX-101) and five were subsequently randomized to lurasidone (ketamine → lurasidone).

### Adherence to investigational product

During the time that participants were enrolled in the clinical trial, adherence to oral doses of investigational product was documented via the AiCure telemedicine system. When doses were missed, study coordinators followed up with the patient to encourage adherence. The adherence identified via electronic monitoring was consistent with the blood levels of investigational drug seen on PK data (below).

### Efficacy Results

The Stage 1 efficacy analyses demonstrated that both ketamine (n=17) and saline (n=5) were associated with a significant reduction in suicidal ideation at 24 and 48 hours post-infusion (primary efficacy endpoint) and improved BDM, CGI-SS, and C-SSRS from pre-infusion baseline in the intention to treat cohort. There were no obvious differences between ketamine and saline treatment groups. Thus, the 48 hour period of Emergency Department admission was associated with remission from depression and suicidality in all subjects.

Primary and secondary endpoints for stage 2 are illustrated in figure 2. For the Stage 2 ITT22 population, the primary outcome measure was change in BDM between baseline and day 42. MMRM regression documented that ketamine → NRX-101 treatment was associated with significantly lower BDM scores compared to ketamine → lurasidone treatment through Days 28 and 42 (p=0.0402 and p=0.0324, respectively) after LOCF analysis. Without LOCF, the difference was significant at Day 28 (p=0.0425) and consistent at Day 42 (p=0.0908). No differences were observed when stratifying for Stage 1 treatment (Table 2). The 7.7 mean difference between treatments for the change from baseline to day 42 has an 1.19 effect size (95% CI: 0.38, 2.00). The effect size for CGI suicidality was 1.38 (95% CI: 0.27, 2.49) was comparable.

**Figure 2:**
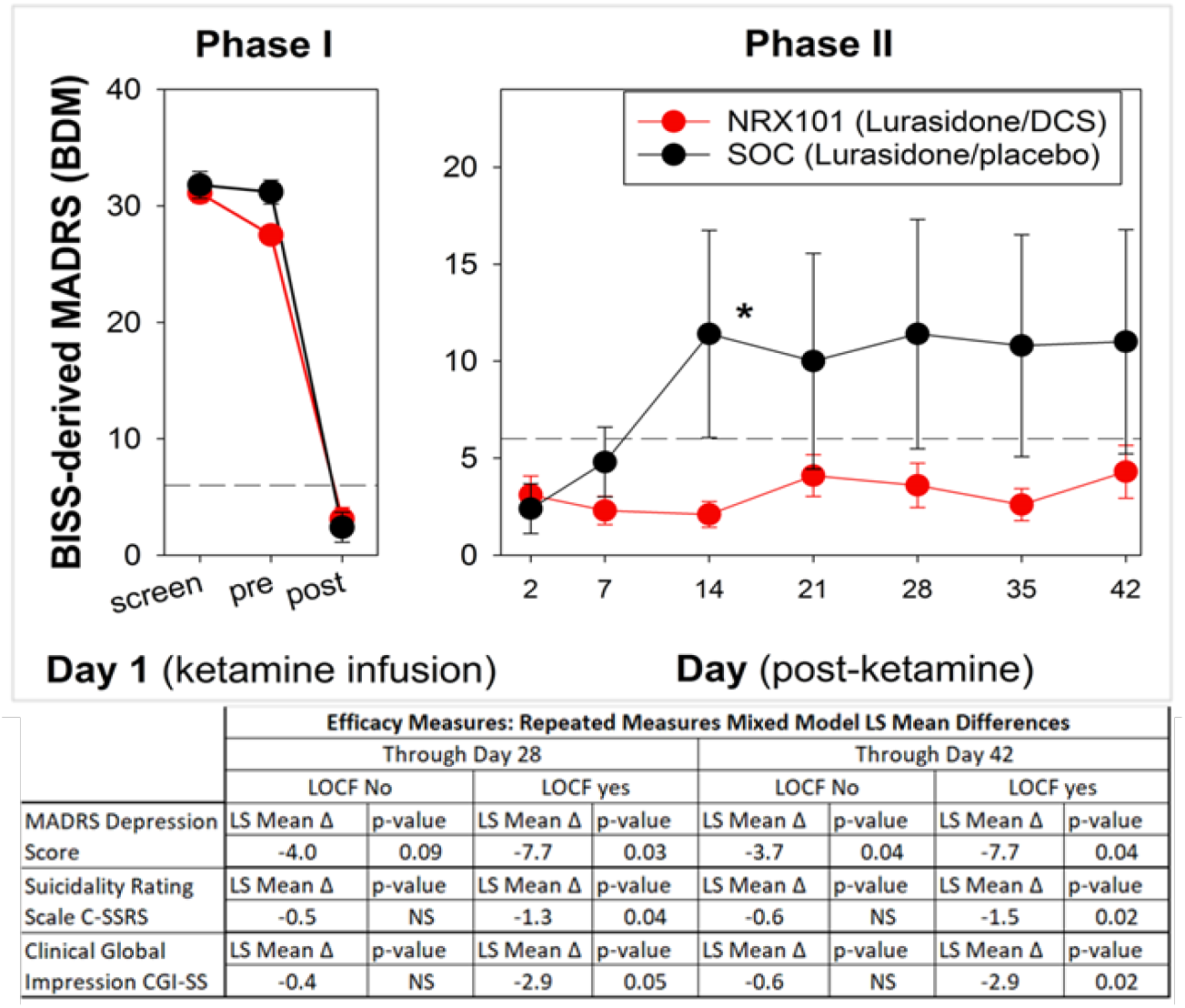
(Graph) Primary Endpoint for those infused with ketamine vs. placebo in phase I (n=22) and those who responded to ketamine in phase I and were randomized in phase II to receive either NRX-101 or lurasidone. Table below depicts primary and secondary endpoints for those randomized in phase II (n=17).

**Table 2 -.** Stage 2 BDM Response Profile Mixed Model Repeated Measures.

| Populations | Day | lurasidone<br>(N=5) | NRX-101<br>(N=12) | NRX-101 -<br>lurasidone<br>(N=17) |
| --- | --- | --- | --- | --- |
|  |  | LSMean | LSMean | LSDifference (p-value) |
| No LOCF | 2 | 2.4 | 3.0 | 0.6 (0.83) |
|  | 7 | 4.6 | 1.7 | -2.9 (0.36) |
|  | 14 | 12.9 | 2.0 | -10.9 (0.002) |
|  | 21 | 5.7 | 5.5 | -0.2 (0.95) |
|  | 28 | 7.7 | 4.0 | -3.7 (0.30) |
|  | BDM 10 Response Profile p-value |  |  | 0.042 |
|  | 35 | 7.3 | 3.3 | -4.0 (0.29) |
|  | 42 | 7.7 | 4.9 | -2.8 (0.46) |
|  | BDM 10 Response Profile p-value |  |  | 0.09 |
| LOCF | 2 | 2.4 | 3.0 | 0.6 (0.85) |
|  | 7 | 4.8 | 2.0 | -2.8 (0.39) |
|  | 14 | 11.4 | 2.2 | -9.2 (0.006) |
|  | 21 | 10.0 | 4.5 | -5.5 (0.09) |
|  | 28 | 11.2 | 3.5 | -7.7 (0.02) |
|  | BDM 10 Response Profile p-value |  |  | 0.04 |
|  | 35 | 10.8 | 2.8 | -8.0 (0.02) |
|  | 42 | 11.0 | 3.7 | -7.3 (0.04) |
|  | BDM 10 Response Profile p-value |  |  | 0.03 |

BDM = BISS-derived MADRS score; ITT1 = Intent-to-Treat Stage 1 Population; ITT2 = Intent-to-Treat Stage 2 Population; LOCF = Last Observation Carried Forward

The key secondary efficacy endpoint for Stage 1 was the difference in mean change in the BDM from pre-infusion baseline to final observation (Day 1 or Day 2 post-infusion) between ketamine and saline treatment groups. For the Stage 1 ITT22 population, the mean baseline BDM score reported at pre-infusion was 29.6 (range: 12-38; ketamine, mean: 28.9, range: 12-38; saline, mean: 31.8, range: 29-37). Both ketamine and saline treatment statistically significantly decreased the mean BDM score at Visit 1 (ketamine, mean: 2.6, range: 0-8, p<0.0001; saline, mean: 5.6, range: 0-19, p=0.0033) and at the end of Stage 1 (ketamine, mean 3.3, range: 0-9, p<0.0001; saline, 5.6, range: 0-19, p=0.0033). There were no statistically significant differences in the mean BDM score between ketamine and saline treatment groups. Neither ketamine nor saline treatment significantly changed the mean BPRS+ score.

The updated key secondary efficacy endpoint for Stage 2 was the difference in the relapse rate between DCS plus lurasidone and lurasidone treatment groups. There was no significant difference observed after stratifying the Stage 2 ITT22 population by its Stage 1 saline treatment (Saline → DCS plus lurasidone and Saline → lurasidone, p=1.000). There was, however, a numerical trend towards a decrease in the relapse rate of the ketamine → NRX-101 group (0%, 0/12 patients) compared to that of the ketamine → lurasidone group (40%, 2/5 patients) (p=0.0735).

The Clinical Global Impression – Suicidality Scale (CGI-SS) was both a secondary efficacy measure and a safety measure in this study. CGI-SS was ascertained by the study physician who was blinded to treatment assignment and blinded to the assessments of the independent raters. The Stage 1 pre-infusion baseline mean CGI-SS total score of the ITT22 population was 17.4 (range: 10-20; ketamine, mean: 17.0, range: 10-20; saline, mean: 18.6, range: 14-20). Both ketamine and saline treatment statistically significantly decreased the mean CGI-SS score from pre-infusion baseline to Visit 1 (ketamine, mean: 2.1, range: 0-9, p<0.0001; saline mean: 1.2, range: 0-4, p=0.0009) and End of Stage 1 (ketamine, mean: 2.1, range: 0-9, p<0.0001; saline, mean: 1.2, range: 0-4, p=0.0009).There were no statistically significant differences observed between ketamine- and saline-treated patients.

MMRM regression was used to interpret changes in CGI-SS for Stage 2 data both without and with LOCF. For the Stage 2 ITT22 population, ketamine → NRX-101 treatment significantly lowered CGI-SS Response Profiles compared to ketamine → lurasidone treatment through Day 28 and Day 42 when imputing LOCF (p=0.0457 and p=0.0186, respectively), but not without LOCF (p=0.1485 and p=0.1527, respectively). No differences were observed in the CGI-SS Response Profiles of MMRM by Stage 1 treatment through Days 28 and 42. With LOCF analyzed, these data suggest that ketamine followed by NRX-101 treatment is significantly associated with a lower (i.e., better) rating of the subject’s suicidality by the treating physician compared to ketamine → lurasidone treatment.

The pre-infusion baseline mean C-SSRS score (suicidal ideation) for the Stage 1 ITT22 population was 4.7 (range: 1-5; ketamine, mean: 4.6, range: 1-5; saline, mean: 5.0, range: 5-5). Both ketamine and saline treatment statistically significantly decreased the mean C-SSRS scores from pre-infusion baseline to Visit 1–4 hours (ketamine, mean: 0.3, range: 0-5, p<0.0001; saline, mean: 0.6, range: 0-2, p=0.0004), Visit 1–10 hours (ketamine, mean: 0.1, range: 0-1, p<0.0001; saline, mean: 0.2, range: 0-1, p<0.0001), Visit 1–24 hours (ketamine, mean: 0, range: 0-0, p<0.0001; saline, mean: 0.2, range: 0-1, p<0.0001), and End of Stage 1 (ketamine, mean: 0, range: 0-0, p<0.0001; Saline, mean: 0.2, range: 0-1, p<0.0001). There was no statistically significant difference observed between ketamine and saline treatment groups.

For the Stage 2, MMRM regression was performed first without LOCF then repeated with LOCF. For the ITT22 population, ketamine → NRX-101treatment significantly lowered C-SSRS (ideation) Response Profiles compared to ketamine → lurasidone treatment through Day 28 and Day 42 when imputing LOCF (p=0.0419 and p=0.0210, respectively) but not without LOCF through Days 28 and 42 (p=0.1125 and p=0.1112, respectively). No difference was observed in the C-SSRS Response Profiles after stratifying by Stage 1 treatment through Days 28 and 42.

### Safety Results (See Supplementary Materials)

In the Stage 1 Safety population, 11 of the 17 ketamine-treated patients (64.7%) experienced a total of 29 AEs whereas no subject treated with saline (0%) experienced an AE. None of these 29 AEs was considered severe and no SAE was recorded. Eleven (68.8%) DCS plus lurasidone-treated patients in the Stage 2 Safety population experienced 36 total AEs, none of which was considered severe and no SAE was experienced. Overall, three AEs were considered drug-related (blurred vision, sedation, and vulvovaginal candidiasis) and 19 were considered possibly related. As a result of these AEs, 12 dose reductions and 3 dose increases of DCS plus lurasidone were made.

Overall, no severe AEs, SAEs, or deaths were experienced in patients treated with ketamine → NRX-101. Three subjects in the ketamine → lurasidone group experienced SAEs (angina pectoris, suicidal ideation, and wound). The angina pectoris, which resulted in hospital admission was later deemed to be non-physiologic in nature and the patient was diagnosed with depression and transferred to inpatient psychiatric care.

The trial was not powered to prove a statistically significant difference on BARS akathisia scores. However, the study demonstrates a 1.0 point increase from baseline in the lurasidone group and a -0.2 decrease in BARS score in the NRX-101 group (d= 1.1; t-test on difference; P=.14). This trend is consistent with the preclinical data and suggests a meaningful effect in reduction of lurasidone-induced akathisia might be seen in an adequately powered trial. The anticipated sample size of the phase 2 trial has >90% power to test the difference on BARS that was seen in STABIL-B.

No mean differences were observed between the NRX-101 and lurasidone groups on the Patient Reported Inventory of Side Effects (PRISE), although some differences were noted on specific items (see supplementary materials).

### Pharmacokinetics Results (see supplementary materials)

DCS and lurasidone blood concentrations were recorded for the Stage 2 Safety populations. The blood levels obtained were consistent with oral ingestion of the investigational product.

## DISCUSSION

The results of this Phase 2b/3 study in 22 patients demonstrate that a sequential treatment of a single infusion of ketamine followed by NRX-101, a fixed dose combination of DCS and lurasidone, is significantly more efficacious than ketamine followed by lurasidone at maintaining reduction in depression (p=0.03) and suicidal ideation, as measured by C-SSRS (p=0.02) and by CGI-SS (p=0.03). There is a trend toward a reduction in relapse (p=0.07). However, ketamine treatment alone did not demonstrate superiority in treating acute suicidal ideation and behavior compared to saline, likely because of the substantial ketamine expectation effect that is now well known in the literature.

The statistically significant separation in stage 2 between NRX-101 and lurasidone alone remained significant on MMRM throughout the 42-day time interval, suggesting that NMDAR antagonist agents may exert both an antidepressant effect and a reduction in suicidal ideation. The decrease in suicidal ideation observed with NRX-101 was seen both on C-SSRS and MADRS as assessed by independent raters and on the CGI-SS scale ascertained by the treating site physician, suggesting a consistency in effect. This beneficial effect on suicidality associated with DCS was also reported by Chen (18) in patients with bipolar disorder who were actively suicidal at baseline.

While the sample size in this study was not sufficient to prove a statistically-significant difference in relapse (0/12 v. 2/5; P=.07), the numerical difference observed suggests that a meaningful difference may be seen in an adequately powered trial. Patients in this study were released from the Emergency Department within 2 days and required hospitalization only after relapse.

No significant safety events were observed following infusion of either ketamine or saline placebo. No SAEs occurred in the ketamine → NRX-101 group. However, three SAEs were reported in the ketamine → lurasidone group with two instances of hospitalization for relapse. Therefore, the sequential regimen of a single infusion of ketamine followed by NRX-101 appears to be safe well tolerated.

Nonclinical studies have identified a significant reduction in akathisia when DCS and other NMDAR antagonists are added to lurasidone and other 5-HT2A antagonists. While this trial was underpowered to prove a difference in akathisia between the NRX-101 and lurasidone only groups, a high effect size was seen (d=1.1) with an increase in akathisia seen in the lurasidone group and a decrease in akathisia seen in the NRX-101 cohort at a trend level of significance (P=.14). This effect will be further explored in future trials.

The effect size in excess of 1.0 determined for the primary endpoint in this study is consistent with the effect sizes reported for DCS in treatment resistant depression by Heresco-Levy^24^ and by Chen in patients with bipolar depression^25^. On the basis of these findings, it was determined that a randomized trial enrolling 72 participants would have near 90% power to detect a statistically significant separation between NRX-101 and lurasidone on both the primary endpoint (mean 5 unit delta) and the secondary CGI-SS endpoint (mean 2 unit delta). This sample size would have yield 80% power to detect a 0.67 effect size, which is set as the lower bound for a clinically-significant result.

Based on these findings, the FDA awarded Breakthrough Therapy Designation and granted a Special Protocol Agreement for a Phase 3 registration trial of NRX-101 vs. lurasidone in treatment of patients with Severe Bipolar Depression and Acute Suicidal Ideation or Behavior (NCT03396068) under Breakthrough Therapy Designation with a sample size of 72 patients at 2:1 randomization.

## CONCLUSION

Patients treated with an oral NMDAR antagonist medication (D-cycloserine) as part of a fixed dose combination with lurasidone (NRX-101), a third-generation antipsychotic indicated for treatment of Bipolar Depression following improvement with ketamine, were shown to have significantly lower levels of depression and ASIB, together with a numerical trend towards reduced likelihood of relapse and rehospitalization than were patients treated with lurasidone alone. A numerical trend towards reduced levels of akathisia was seen (P=0.14) in the NRX-101-treated patients, consistent with findings in non-clinical studies. Registration studies under an FDA Special Protocol Agreement and Breakthrough Therapy Designation are now underway.

## Supporting information

Equator (CONSORT) checklist

## Data Availability

All data produced in the present study are available upon reasonable request to the authors

https://clinicaltrials.gov/ct2/show/results/NCT02974010

## Supplementary Materials

### Detailed Safety Results

In the Stage 1 Safety population, all patients received the entire dose of ketamine (0.5 mg/kg, n=17) or saline (n=5). For the Stage 2 safety population, all patients were administered an average of 58.6 oral capsules of DCS plus lurasidone or lurasidone (range 1-88; DCS plus lurasidone, n=16, mean: 59.1 capsules, range 1-88 capsules; lurasidone, n=6, mean: 57.3 capsules, range 6-82 capsules). This led to an average of 30.6 days with at least one dose of a study treatment (range 1-45 days; DCS plus lurasidone, mean: 30.9 days, range: 1-45 days; lurasidone, mean: 40.5 days, range: 3-43 days.

In the Stage 1 Safety population, 11 of the 17 ketamine-treated patients (64.7%) experienced a total of 29 AEs whereas no subject treated with saline (0%) experienced an AE. None of these 29 AEs was considered severe and no SAE was recorded. Sixteen of these 29 AEs were known to be associated with ketamine usage and were considered related to ketamine (two events each of dissociation, dizziness, euphoric mood, hypertension, and hypoesthesia and one event each of blurred vision, diplopia, dysmetropsia, restlessness, tinnitus, and vomiting) and another 12 were considered possibly related to ketamine treatment (Table 4).

Eleven (68.8%) DCS plus lurasidone-treated patients in the Stage 2 Safety population experienced 36 total AEs, none of which was considered severe and no SAE was experienced (Table 4). Overall, three Aes were considered related to (blurred vision, sedation, and vulvovaginal candidiasis) and 19 were considered possibly related to (four events of restlessness, three events each of akathisia and fatigue, and one event each of anorgasmia, blurred vision, ejaculation delayed, headache, hypersomnia, lethargy, sedation, somnolence, and increased weight) DCS plus lurasidone. As a result of these AEs, 12 dose reductions and 3 dose increases of DCS plus lurasidone were made.

Four (66.7%) lurasidone-treated patients in the Stage 2 Safety population experienced 11 total AEs (Table 4), three of these were considered severe (one event of angina pectoris and two events of suicidal ideation) and two patients experienced three SAEs (angina pectoris, suicidal ideation, and wound). However, angina pectoris was later determined to be associated with relapse of depression and the patient was transferred to inpatient psychiatric care. Overall, one AE was considered related to (sedation) lurasidone and one AE was considered possibly related to lurasidone. Nine of the 10 AEs did not result in a dose change and one AE resulted in an unknown change.

Overall, no severe AEs, SAEs, or deaths were experienced in patients treated with ketamine → NRX-101. Three SAEs were reported in the ketamine → lurasidone group: 1 cardiovascular, Mean Patient Rated Inventory of Side Effects (PRISE) and Distressing PRISE did not differ between NRX-101 and lurasidone-treated groups. While ketamine predictably affected blood pressure, systolic blood pressure (SBP) and diastolic blood pressure (DBP) changes were normalized within 200 and 80 minutes post-treatment, respectively. Additionally, mean pulse and weight did not change in ketamine- or DCS plus lurasidone-treated patients compared to pre-infusion baseline, while mean SBP and DBP were unchanged in DCS plus lurasidone-treated patients compared to post-infusion baseline (Table 4).

The STABIL-B was not powered to prove a statistically significant difference on BARS akathisia scores. However, the study demonstrates a 1.0 point increase from baseline in the lurasidone group and a -0.2 decrease in BARS score in the NRX-101 group (d= 1.1; t-test on difference; P=.14). This trend is consistent with the preclinical data and suggests a meaningful effect in reduction of lurasidone-induced akathisia might be seen in an adequately powered trial. The anticipated sample size of the phase 2 trial has >90% power to prove the difference on BARS that was seen in STABIL-B. Therefore, the concept of a sequential treatment of a single infusion of ketamine followed by oral DCS plus lurasidone demonstrated a convincing safety profile and should be considered safe for use treating bipolar depression in patients with ASIB.

### Pharmacokinetics Results

DCS and lurasidone HCl blood concentrations were recorded for the Stage 2 Safety populations (supplemental Table 5). DCS and lurasidone were not detectable before the drug administration. For patients treated with NRX-101 in the Safety population, the mean DCS concentration was 5.709 µg/mL, 17.820 µg/mL, 12.104 µg/mL, and 24.373 µg/mL at Visit 2 post-dose, Visit 7 post-dose, Visit 11 pre-dose, and Visit 11 post-dose, respectively, and the mean lurasidone concentration was 10.366 ng/mL, 10.135 ng/mL, 10.859 ng/mL, and 36.965 ng/mL at Visit 2 post-dose, Visit 7 post-dose, Visit 11 pre-dose, and Visit 11 post-dose, respectively (Table 5). For patients treated with lurasidone, the mean lurasidone concentration was 9.813 ng/mL, 9.688 ng/mL, 7.227 ng/mL, and 27.750 ng/mL at Visit 2 post-dose, Visit 7 post-dose, Visit 11 pre-dose, and Visit 11 post-dose, respectively (Table 5).

## Notes

### Competing Interest Statement

DCJ,JCJ, RBE receive compensation from and have equity interest in NRx Pharmaceuticals, Inc. Lavin Statistical Associates is paid for independent statistical analysis by NRx Pharmaceuticals, Inc.

### Clinical Trial

www.clinicaltrials.gov NCT 02974010

### Clinical Protocols

https://clinicaltrials.gov/ProvidedDocs/10/NCT02974010/Prot_SAP_000.pdf

### Funding Statement

This study was funded by NRx Pharmaceuticals, Inc. (Wilmington, DE)

### Author Declarations

Advarra IRB (Cincinnati, OH) gave ethical approval for this work. Approval Number: 201704604 The research was conducted under an IND 129194 granted by the US FDA. The study was under the supervision of an independent Data Safety Monitoring Board.

